# Non-alcoholic fatty liver disease (NAFLD) prevalence in Latin American and the Caribbean: Protocol for a systematic review

**DOI:** 10.1101/2020.11.16.20232744

**Authors:** Yesmi Arcelia Ortega Rojas, Claudia Lucía Vidal Cuéllar, Karina Mercedes Aparicio Barrón, Rodrigo Carrillo Larco

**Affiliations:** School of Medicine, Universidad Peruana Cayetano Heredia, Lima, Perú; CRONICAS Centre of Excellence in Chronic Diseases, Universidad Peruana Cayetano Heredia; Department of Epidemiology and Biostatistics, Imperial College of London, UK

**Keywords:** Non alcoholic fatty liver disease, non-alcoholic fatty liver disease, NAFLD

## Abstract

**Background:** Non-alcoholic fatty liver disease (NAFLD) is the most common worldwide cause of chronic liver disease and it carries a significant economic burden for the healthcare system. The worldwide prevalence of NAFLD appears to be 25%, with the highest rates in Latin America and the Caribbean (LAC) and the middle west. Prevalence of NAFLD in LAC has not been well represented in multiple global meta-analyses because they did not include regional search engines or excluded papers in Spanish. Furthermore, older estimates may not represent the current epidemiological situation given the recent rise of several risk factors like obesity, unhealthy diet and lifestyles in LAC.

**Objective:** To estimate the prevalence of NAFLD in the general adult population of LAC.

**Methods:** Systematic review and meta-analysis. We will conduct a search in OVID (MEDLINE, Embase, Global Health), Cochrane Library, and LILACS. Search terms include those related to non-alcoholic fatty liver disease, along with countries in LAC. We seek observational studies with a random sample of the general population, closed populations, and patients; results will be presented for each of these groups. Titles and abstracts will be screened by two reviewers independently. After the screening phase, we will download the full-text of the selected publications and two reviewers will comprehensively read these papers. If there is any discrepancy in any of these two selection phases, it will be solved by consensus by the two reviewers or by a third party. An extraction data form in a spreadsheet will be developed and tested with a random sample of five selected studies. The data extracted will include study details, sample size, method of NAFLD diagnosis and prevalence of NAFLD. Data extraction will be conducted by two reviewers independently. We will use the risk of bias tool proposed by Hoy and colleagues. We will report the main features of the selected reports in tables, this will include the prevalence estimates. We will pool the prevalence estimates. We will only conduct the meta-analysis if there are three or more individual estimates and there is not large heterogeneity amongst them. We will conduct all statistical analyses using Stata 16.0

**Conclusions:** This systematic review and meta-analysis will provide updated evidence about the prevalence of NAFLD in LAC. Thereby, we will increase NAFLD awareness as a public health problem in LAC, and our results can inform strategies and interventions to reduce NAFLD burden in LAC. We will also pinpoint research needs and discuss future steps so that more robust evidence about NAFLD becomes available in LAC.

## BACKGROUND

Non-alcoholic fatty liver disease (NAFLD) is the most common worldwide cause of chronic liver disease and it represents a wide spectrum of conditions, including liver cirrhosis and hepatocellular carcinoma in advanced stages.^1, 2^ NAFLD is consider a multisystemic disease associated with high morbidity and mortality due to extrahepatic conditions such as Type 2 Diabetes Mellitus (T2DM), Cardiovascular Diseases (CVD), Chronic Kidney Disease (CKD) and metabolic syndrome.^4, 5, 12^ Furthermore, NAFLD carries a significant economic burden for the healthcare system estimated at 1,613 dollars per patient per year in the USA and 35 billions euros per year per country in Europe.^6^ A projection estimates that the economic burden of NAFLD would increase to 1.005 trillions dollars per year in the USA and to 334 billions euros in Europe, if NAFLD has a similar growth curve as obesity.^6^ Overall, the burden of NAFLD at the patient and population level is nontrivial.

The worldwide prevalence of NAFLD appears to be 25%, with the highest rates in South America (Brazil, Chile, Colombia) and the middle west.^3^ The prevalence estimates have increased in the last decade due to numerous factors like unhealthy diet and lifestyles changes and an increase in obesity prevalence.^6^ The high prevalence of NAFLD in Latin America and the Caribbean (LAC) is also associated with genetic predisposition, ethnicity, low physical activity, alcohol consumption, high prevalence of metabolic syndrome as well as T2DM, and limited access to the healthcare system.^6, 7^ Not only are people in LAC at increased risk of NAFLD because of high rates of these risk factors, but it seems that LAC populations might experience worse cases of NAFLD because of a genetic polymorphism associated with steatosis, steatohepatitis and fibrosis that is present more often in Hispanic populations than in African Americans.^7, 13^ With already large prevalence estimates, high rates of risk factors, and apparently worse NAFLD presentation, LAC needs solid epidemiological evidence about NAFLD to inform public health interventions and research needs; nevertheless, NAFLD prevalence estimates in LAC are not current and lack in several countries of this region.

NAFLD prevalence estimates have been summarised by recent meta-analysis based on studies conducted over a decade ago.^15^ It is important to generate updated evidence about the prevalence of NAFLD in LAC, because older estimates may not represent the current epidemiological situation given the recent rise of several risk factors like obesity, unhealthy diet and lifestyles in LAC.^7, 8, 9, 14^ In addition to the dearth of recent studies, NAFLD prevalence estimates from LAC - and other low- and middle-income countries - have not been well represented in multiple global meta-analyses,^3, 16^ because they did not include regional search engines or excluded papers in spanish.^6, 8^

Due to the great impact of NAFLD on patients with this condition as well as the large burden that NAFLD poses on the healthcare system and the economy, we aim to provide solid and updated evidence of the NAFLD prevalence in LAC. Thereby, we will increase NAFLD awareness as a public health problem in LAC, and our results can inform strategies and interventions to reduce NAFLD burden in LAC. We will also pinpoint research needs and discuss future steps so that more robust evidence about NAFLD becomes available in LAC.

## METHODS

### Objectives

To estimate the prevalence of NAFLD in the general adult population of LAC.

### Study design

Systematic review. We will follow the PRISMA guidelines for systematic reviews and meta-analyses.^10^

### Eligibility criteria

#### PICO question

What is the prevalence of NAFLD in the general population of adults in LAC?

We will include studies that fulfill the following criteria:

- **Population:** Adult individuals (18+ years old) selected from the general population; captive or closed populations; or patients from any healthcare facility. Participants should have been selected following any sort of random sampling technique; in other words, we will not include purposive or non-probabilistic samples. Participants should be natural and residents of a country in LAC.
- **Intervention:** Not applicable
- **Comparison:** Not applicable
- **Outcome:** Prevalence of NAFLD diagnosis as defined by the original study; the diagnosis could have been based on laboratory analysis (e.g., blood samples), imaging (e.g., tomography or ultrasonography) and/or invasive methods (e.g., liver biopsy). That is, when the NAFLD diagnosis was based on self-reported information alone (e.g., questionnaires), we will not include this original report.

The study designs that will be included are observational studies: population-based surveys, cross-section and cohort (baseline) epidemiological studies. If there were systematic reviews on this subject, we will revise the reference lists to identify relevant original sources.

We will include studies even if these did not exclude other causes of liver disease (e.g., viral infections), did not screen for excess alcohol consumption, or did not exclude people consuming hepatotoxic medications. Nevertheless, we will take these characteristics in consideration during the data extraction and analysis (e.g., to stratify results).

### Exclusion criteria

The following study designs will be excluded: case reports, case series, letters to the editor, editorial, narrative reviews, clinical trials, case-control and systematic reviews.

### Literature search and data collection

We will search in OVID including MEDLINE, EMBASE and Global Health; we will also search the Cochrane Library and LILACS. We will use the search terms detailed in the Search Strategy (Table 1).

**Table 1.**
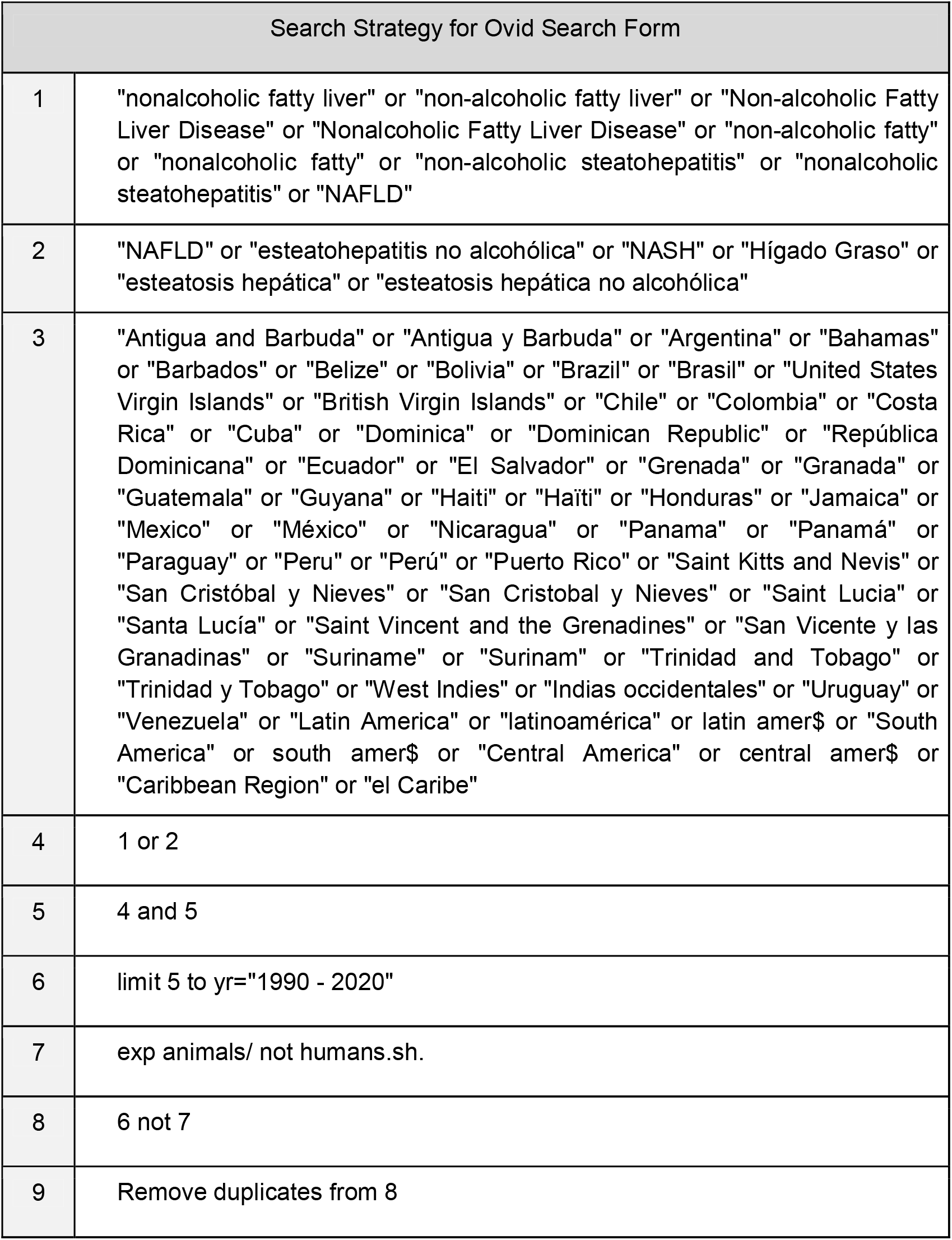

The search will be restricted to human subjects only; no language restriction will be implemented. We will restrict the search to results published between 1990 and 2020.

First, titles and abstracts will be screened by two reviewers independently, looking for studies that meet the selection criteria above detailed. After the screening phase, we will download the full-text of the selected publications and two reviewers will comprehensively read these papers. If there is any discrepancy in any of these two selection phases, it will be solved by consensus by the two reviewers or by a third party. Finally, selected studies will be checked to identify whether the same data were used in more than one publication; if so, we will choose the publication with more information relevant to our research question, or the publication with the largest sample size. We will record the reasons for exclusion during the second selection phase (full-text). The number of included/excluded publications across the selection phases will be detailed in a flow diagram according to the PRISMA recommendations.

### Data collation

An extraction data form in a spreadsheet will be developed and tested with a random sample of five selected studies. If need be, the extraction form will be updated and will not be modified after data collation started. Two reviewers, independently, will carry out data collation. Disagreements will be resolved by consensus or by a third party.

The data extracted will include:

- Study details: First author, corresponding author, article title, country, year of publication and year of data collection.
- Sample size: Number of participants, sampling procedure, characteristics of the study population (overall mean age and proportion of male subjects), inclusion criteria and exclusion criteria, and whether the study is nationally representative. We will also record whether the study population was tested for relevant viral infections, binge alcohol drinking and consumption of hepatotoxic medications.
- Study methodology: Method of NAFLD diagnosis.
- Results: Prevalence of NAFLD in adults from the general population, captive or closed populations or patients. If available, prevalence estimates will be extracted by gender and/or age group. Prevalence estimates will be extracted along with 95% confidence intervals.

If we find any publication in which the methodology or results are unclear, we will try to contact the corresponding author through email. We will wait for two weeks, in case we do not have an answer from the corresponding author and doubts could not be solved by other means, the original publication will be excluded.

### Risk of bias and methodological quality of studies

Each selected report will be subject to risk of bias assessment. We will use the risk of bias tool proposed by Hoy and colleagues.^11^ If during this risk of bias assessment there were any discrepancies, these will be solved by consensus between reviewers or by a third party.

### Statistical analysis

Following a qualitative summary approach, we will report the main features of the selected reports in tables, this will include the prevalence estimates. If relevant, we will summarise the evidence in figures showing the prevalence estimates by country and/or year of data collation. Following a quantitative summary approach, we will pool the prevalence estimates. We will only conduct the meta-analysis if there are three or more individual estimates and there is not large heterogeneity amongst them. We will follow a random-effects meta-analysis with the DerSimonian and Laird method. We will present pooled prevalence estimates along with a metric of heterogeneity (I^2^). If possible, i.e., if three or more estimates are available, pooled estimates will also be presented by gender. We will conduct all statistical analyses using Stata 16.0 (College Station, Texas 77845 USA).

## Data Availability

This is a systematic review protocol, and no data have been analysed.

## Conflicts of interest

All authors declare to have no conflicts of interest.

## Funding

Rodrigo M Carrillo-Larco is supported by a Welcome Trust International Training Fellowship (214185/Z/18/Z)

## Notes

### Competing Interest Statement

The authors have declared no competing interest.

### Funding Statement

Rodrigo M Carrillo-Larco is supported by a Wellcome Trust International Training Fellowship (214185/Z/18/Z).

### Author Declarations

This is a systematic review protocol. No human subjects were involved in this project. Thus, this study was classified as non-human subject research. Therefore, no approval was needed from an IRB/ethics committee

